# Mapping Inequities in Global Vaccine Sentiment Research

**DOI:** 10.1101/2025.01.11.25320376

**Authors:** Duilio Balsamo, Vittoria Offeddu, Zhina Aghamohammadi, Chiara Chiavenna, Laura P. Leone, Elena D’Agnese, Deepak Sharma, Aleksandra Torbica, Soheil Shayegh, Javier Andreu-Perez, Alessia Melegaro

## Abstract

**Introduction:** Negative public sentiment towards vaccination (PSV) poses significant challenges to the effectiveness of immunization programs, with dramatic effects on morbidity and mortality for vaccine-preventable diseases. Yet, health research is often shaped by economic and geopolitical factors rather than countries’ epidemiological or healthcare needs. This study examines global patterns and drivers of PSV research on five vaccines –polio, measles, human papillomavirus, influenza, and SARS-CoV-2– and evaluates the COVID-19 pandemic’s impact on research volume, focus, and distribution.

**Methods:** We conducted a machine-learning-assisted literature search on PSV without geographical, language or time constraints. Using natural language processing, network and statistical analyses, we examined the global PSV research landscape and identified geographical, epidemiological, and economic drivers.

**Results:** We analysed 13,287 articles and detected consistent literature growth from 1980 onwards, with vaccine-specific peaks following key licensing events. After 2020, publication volumes rose above projections for influenza (32%; 95%CI = 20%, 46%) but declined for polio (–56%; 95%CI = –68%, –26%) and measles (–17%; 95%CI = –33%, 9%). Although PSV research had global coverage, its distribution was markedly imbalanced, largely shaped by country-specific economic factors. A few high-income countries (HIC) produced 72% of publications and the likelihood of a country being studied varied by income and vaccine. Foreign authorship also increased as the income of the studied country decreased (over 75% in LMIC vs. below 50% in HIC).

**Conclusion:** PSV research reveals persistent inequities, with a misalignment between countries leading research and populations most in need of its outcomes. These inequities, further exacerbated by COVID-19 disruptions, reflect systematic imbalances in global health. Our findings underscore the need to decolonise research by fostering leadership, agenda-setting, and accountability that centres on the necessities of affected communities. Achieving this will require funding and publication reforms that promote equitable collaborations and elevate local priorities alongside long-standing global health objectives.

**Key messages:** *What is already known on this topic:* Public sentiment towards vaccination (PSV) significantly influences vaccination campaign outcomes. While bibliometric studies have explored PSV research patterns, economic and contextual drivers in this research field remain underexplored, and international collaboration dynamics lack in-depth analysis.

*What this study adds:* This study identifies significant economic disparities shaping PSV research, with high-income countries dominating the field and directing studies toward low-income settings with limited local author representation. It also highlights the COVID-19 pandemic’s role in shifting research focus from critical childhood vaccines to SARS-CoV-2, exacerbating these imbalances.

*How this study might affect research, practice or policy:* By revealing inequities in PSV research, this work underscores the need for more equitable and inclusive research initiatives, prioritizing public health needs while addressing systemic disparities in global health research and policy.

## Introduction

The primary goal of global health research is to promote health equity and strengthen healthcare systems by bridging global knowledge with local interventions.^1^ However, global health initiatives are heavily influenced by economic factors.^2^ Researchers from low– and middle –income countries (LMIC) face limited access to global health research grants,^3^ and are underrepresented in peer-reviewed publications.^4^ Furthermore, 90% of all health research funding targets health issues of industrialized countries,^5^ marginalizing lower-income regions, where health burdens are typically greater. This perpetuates inequities, with researchers from high-income countries (HIC) dominating research activities also in LMIC, resulting in inadequate research agendas that fail to meet local public health needs.^2^ Vaccine uptake and childhood immunization patterns across the globe represent a significant example of this dynamic.

Despite vaccines’ efficacy, global uptake remains suboptimal in most countries, pockets of zero-dose individuals persist,^6^ and public sentiment towards vaccination (PSV) is imbued with widespread hesitancy.^7^ Over time, considerable research efforts have been directed towards investigating the intricate mechanisms shaping PSV, resulting in a heterogeneous, multi-disciplinary, and fast-growing body of literature.^8^ Numerous reviews attempted to systematize and summarize the existing evidence on PSV.^9^ However, due to the sheer amount and complexity of the published data, this task is proving increasingly difficult, especially without the aid of machine-assisted methods. Identifying the main drivers of PSV research might facilitate the definition of future research priorities and provide an evidence-based rationale to support a more appropriate allocation of resources.

To what extent PSV research is shaped by the interplay between economic factors, local disease endemicity, and the effective need to study PSV locally is not fully understood,^8,10–12^ especially in light of disruptive events, such as the COVID-19 pandemic. The exceptional circumstances in which the SARS-CoV-2 vaccine was licensed, together with the determining role it has played in curbing the pandemic, significantly intensified research efforts and public discussions on PSV.^13^ However, the pandemic also diverted a significant portion of global academic efforts and funding from other research to COVID-19-related contributions, which benefitted from fast publication tracks and great academic attention, putting research on other diseases and vaccines at considerable disadvantage.^14–16^

In this work, we investigated the global patterns and drivers of scientific research on PSV, including studies on public attitudes, beliefs, emotions, trust, hesitancy and uptake. To enable a fine-grained yet broad perspective on the literature, we used machine-learning-assisted article selection to perform a large-scale literature search of peer-reviewed publications. We screened for a selected set of vaccines *i.e.*, those against polio, measles, Human PapillomaVirus (HPV), influenza, and SARS-CoV-2, chosen to be representative of different target populations, licensing and distribution histories, and viral pathogens with distinct transmission routes and epidemiology. Using a combination of Natural Language Processing and Large Language Model techniques we enriched each record with standardised geographic metadata about authorship and place of study, and linked each record with epidemiological and economic country-specific metadata. We analysed publication trends for all vaccines, and by benchmarking publication volumes before and after 2020, we quantified the extent to which the COVID-19 pandemic has altered pre-existing publication dynamics. Then, leveraging network and statistical tools, we modelled and analysed the geographical, epidemiological, and economic factors driving scientific focus and academic contribution in the PSV research field. The integrated perspective introduced in this work charts the overall structure of PSV literature at large scale, allowing for direct comparisons across a set of vaccines with distinct characteristics and public attention patterns.

Our paper represents a significant contribution to the literature both in terms of methodology as well as implications for academic research and global health policy. By combining novel techniques with traditional statistical approaches, we offer a strong example of how diverse methods can be jointly applied to address complex research questions. By outlining the structural inequities that influence PSV research agendas, our findings provide important insights to guide future research priorities, with the ultimate goal of a more equitable, balanced, and resilient global health research agenda.

## Methods

### Database construction

To retrieve scientific publications on PSV, we developed a comprehensive search string according to validated guidelines,^17^ incorporating keywords regarding public knowledge, attitudes, awareness, emotions and beliefs related to vaccination, vaccine hesitancy and refusal, trust, and uptake <u>(Table S1)</u>. To optimize the specificity of our search, we limited vaccine-related keywords to the article title, and other search terms to the title and abstract. We refined our search string through subsequent search cycles and qualitative screening of the first n=100 delivered papers, iteratively removing 1) keywords retrieving irrelevant literature, *e.g*. “vaccinia”, and 2) individual terms that were likely to occur together with a synonymous search term in true positive cases and retrieving irrelevant literature otherwise. The literature search was conducted in the MEDLINE and Web of Science Core Collection databases until December 2023, without geographical, language or other time constraints. We chose this database combination to cover a broad spectrum of scientific disciplines while obtaining the best possible balance between sensitivity and specificity.^18^ We removed duplicates using the article DOIs and titles and excluded papers not mentioning any of the vaccines of interest in their titles or abstracts.

Given the large number of retrieved articles, we employed supervised Machine Learning (ML) techniques to identify studies directly related to PSV. We began by manually annotating a sample of 1,000 articles by title and abstract, following a predefined set of inclusion criteria. Articles were retained if they addressed at least one dimension of vaccine sentiment, such as studies on knowledge, attitudes, and practices, or assessed determinants of vaccine acceptance, uptake, or timeliness. Eligible study types included both quantitative and qualitative primary research, secondary data analyses, theoretical frameworks, mathematical models exploring vaccination determinants as well as reviews and editorials. After the first round of screening, performed by two independent reviewers, we obtained an inter-rater agreement *Cohen’s k = 0.8*, indicating substantial agreement. A third reviewer resolved the disagreements, resulting in 376 articles labelled as directly related to PSV and 624 as unrelated.

Next, using this annotated sample, we trained multiple ML classifiers, fine-tuning the pipelines’ hyperparameters in a 5-fold cross-validation setting to maximize classification performance. Model evaluation was conducted on a held-out test set (30% of the annotated data), achieving high performance across models, with all classifiers reaching an F1-score > 0.86 and a Matthews Correlation Coefficient (MCC) > 0.78.^19^ The best-performing model – *Random Forest Classifier* with *bag-of-words* and *tf-idf* preprocessing-achieved an *F1-score of 0.89*, and *MCC of 0.82* on the test set. This model was then applied to classify the full dataset. For each retained article, we identified the *primary vaccine* studied, defined as the most frequently mentioned vaccine in the title and/or abstract, as well as the other mentioned vaccines. If multiple vaccines were referenced equally, the first vaccine appearing in the text was assigned as the primary one.

### Data pre-processing and enrichment

Using OpenAI’s GPT (<u>Supplement-Methodological details</u>) we enriched our corpus by extracting from titles and abstracts of collected articles the *authors’ countries*, *i.e.* the countries of the authors’ institutions, and the *studied countries*, *i.e.,* the countries of the analysis. Each country was then assigned a list of categorical attributes, including income level as of 2021,^20^ time-dependent vaccine-specific coverage, ^21,22^ and time-dependent disease incidence for each related disease.^23,24^ Country income was chosen as a proxy for a country’s research capacity since research institutions, such as universities and public health organisations, largely rely on national funding and infrastructure to conduct their projects. Disease incidence dynamically captures the epidemiological pressure that a country is facing. Together with varying levels of vaccination coverage, these factors are likely to influence the scientific –and public-concern about a health threat, potentially fuelling or slowing down research. All attributes were categorized into four groups (L=low; LM=lower-middle; UM=upper-middle; H=high) based on the empirical quantiles (<u>Supplement-Methodological details</u>). Polio incidence was classified as either “No cases” or “Reported cases”. Due to incomplete data on disease incidence and vaccination coverage for HPV and influenza, we limited the analysis including all attributes to measles and polio.

### Time trend analysis

To characterize the publication trends over time, we modelled the volume of published papers for each vaccine from 2010 to 2019 fitting an exponential function. We conducted this analysis starting from 2010 to ensure inclusion of the period following the licensure of both the HPV and 2009 H1N1 pandemic influenza vaccines. The impact of the SARS-CoV-2 pandemic on PSV-related research was assessed by comparing the predicted publication volumes for 2020-2023, based on the fitted exponential models, with the actual volume produced after 2020, calculating the *percentage difference* (<u>Supplement-Methodological details</u>).

### Geographic network construction

We developed a custom network analysis pipeline to map the geographic direction of global research on PSV. In particular, for each 5-year time interval, we created vaccine-specific networks where each node represents a country, and each directed weighted edge, a connection between countries *i* and *j*, was formed proportionally to the number of papers authored or co-authored by institutions from country *i* focusing on country *j.* Articles with multiple authors’ countries and/or studied countries contributed to multiple edges in the network. We further characterized each node *i* by counting the number of 1) *Domestic* studies, papers on country *i* authored by local researchers alone (the weighted self-loops of node *i*); 2) *Inbound* studies, papers on country *i* authored or co-authored by foreign institutions (the weighted in-degree minus the self-loops); 3) *Outbound* studies, papers on any foreign country authored or co-authored by institutions from country *i* (the out-degree minus the self-loops).

### Statistical analysis on research drivers

We employed a regression framework to investigate the drivers of global PSV research. First, to investigate the probability of a country being *studied*, we fitted logistic regression models using a binary dependent variable indicating whether a country had been the object of at least one research paper on a given vaccine (1 if studied, 0 if not) (<u>Supplement-Methodological details</u>). Second, for countries that were *studied*, we analysed which countries’ factors influenced the proportion of papers authored or co-authored by foreign institutions (*i.e., Inbound* / (*Inbound* + *Domestic*)). We achieved that through a Beta regression model, suitable for modelling proportions (<u>Supplement-</u> <u>Methodological details</u>). Independent variables in both models included 1) time interval, classified as pre-pandemic (2010-2014; 2015-2019) and post-pandemic (2020-2023) periods, 2) country income level, 3) country time-varying vaccination coverage level, and 4) country time-varying disease incidence level. All models except the ones for polio (for which not enough data was available) included an interaction term between income and time to test whether the probability of being studied or the proportion of foreign authorship varied over time across income groups (<u>Supplement-Methodological details</u>).

### Ethics approval

This study did not include human participants and is thus exempt from Institutional Review Board approval and the requirement for informed consent.

## Results

### The unfolding of PSV research over time

Our query across two databases yielded 154,552 articles from 1902 to 2023 (Figure S1). We removed duplicates (*43%*) and screened for references to any of the five selected vaccines, retaining 17,587 articles. Of these, 97.5% were written in English, 2.2% in other European languages, and 0.2% in non-European ones. We then used ML-assisted techniques to exclude non-relevant papers (*24%* of articles on the selected vaccines). We retained a total of *13,287* articles (*18%* of the initial yield) for analysis (Figure S1). Of these, 44% focused on SARS-CoV-2 as the *primary vaccine* (n= 5,818), 25% on influenza (n=3,328), 23% on HPV (n=3,078), 7% on measles (n=883), and 1% on polio (n=180).

The volume of PSV literature did not rise significantly until the late 1980s (Figure 1). We observed exponential growth across all vaccines until 2020, with vaccine-specific surges corresponding to the licensing of the HPV and SARS-CoV-2 vaccines, and a volume spike in influenza-related publications following the development of the novel influenza vaccine against the 2009/H1N1 pandemic strain. Using the fitted exponential models (<u>Figure S2</u>), we predicted the volume of publications post-SARS-CoV-2 and compared them against the measured volumes, finding a higher-than-expected increase in the volume of influenza-related research (Perc. Diff. *= 32%; 95%CI = 20%, 46%)*. In contrast, the volume of PSV research on the two routine childhood vaccinations, polio and measles, fell below expectation after the pandemic, with decreases of *Perc. Diff. = –56% (95%CI = –68%, –26%)* and *Perc. Diff. = –17% (95%CI = –33%, 9%)* respectively.

**Figure 1.**
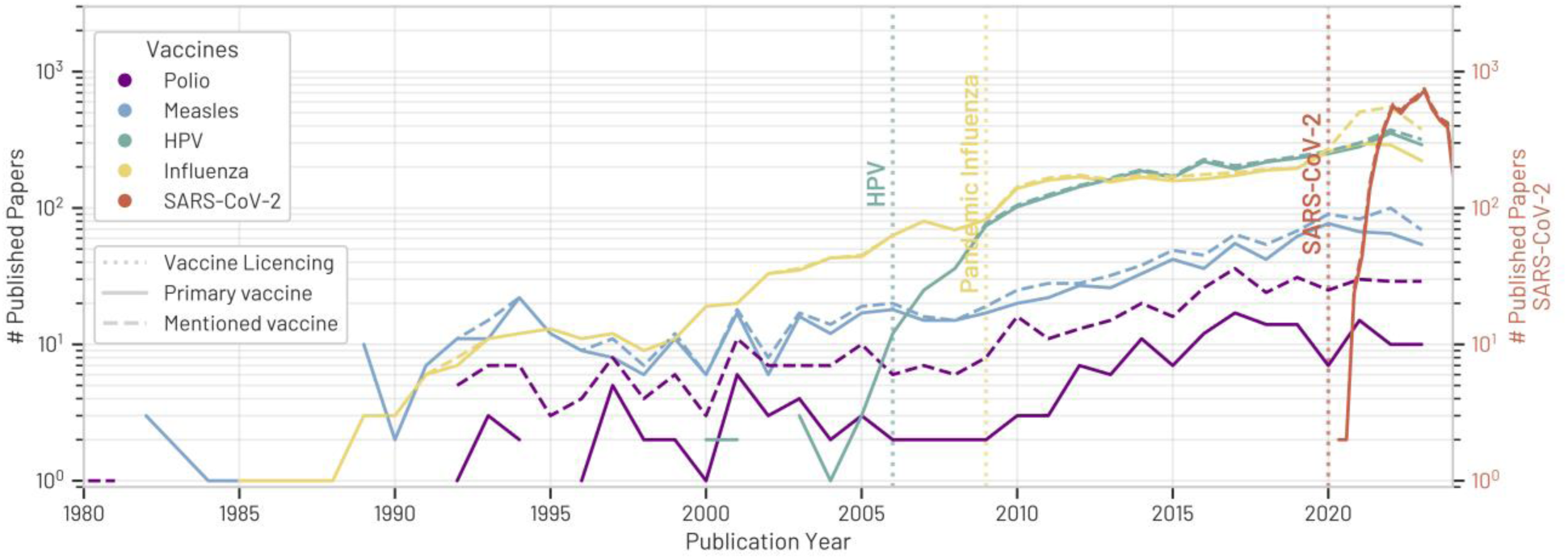
Annual number of published scientific articles on public sentiment towards 5 selected vaccines from 1980 onwards. Dashed lines indicate the total number of articles mentioning each vaccine, reported in different colours, in their title and/or abstract. Solid lines indicate the total number of articles focusing on a specific vaccine as the *primary vaccine*. Vertical lines mark vaccine licensing dates. The licensing date for influenza corresponds to the vaccine against the 2009/H1N1 pandemic strain, while the licensing of the 1945 inactivated seasonal influenza vaccine is omitted. Polio (1955) and measles (1963) vaccine licensing dates precede the emergence of PSV research. For visualization purposes, the number of papers published on SARS-CoV-2 is evaluated and reported quarterly.

Notably, an increasing proportion of papers mentioned more than one vaccine in the text (7·3% before 2020 and 12·9% after 2020). In particular (Figure 2), vaccines sharing similar target populations, such as polio and measles, or those protecting against pathogens with similar characteristics, such as influenza and SARS-CoV-2, two respiratory viruses with pandemic potential, were more frequently studied together. Additionally, several studies on all *primary vaccines* began co-mentioning SARS-CoV-2 during the pandemic. This applied even to vaccines with relatively different characteristics, such as polio, measles, or HPV, suggesting a potential impact of the SARS-CoV-2 pandemic on PSV research. Despite historical trends, the share of articles studying influenza alone also declined after the pandemic, with a large proportion (19·7%) of recent influenza-related articles also referencing the new coronavirus.

**Figure 2.**
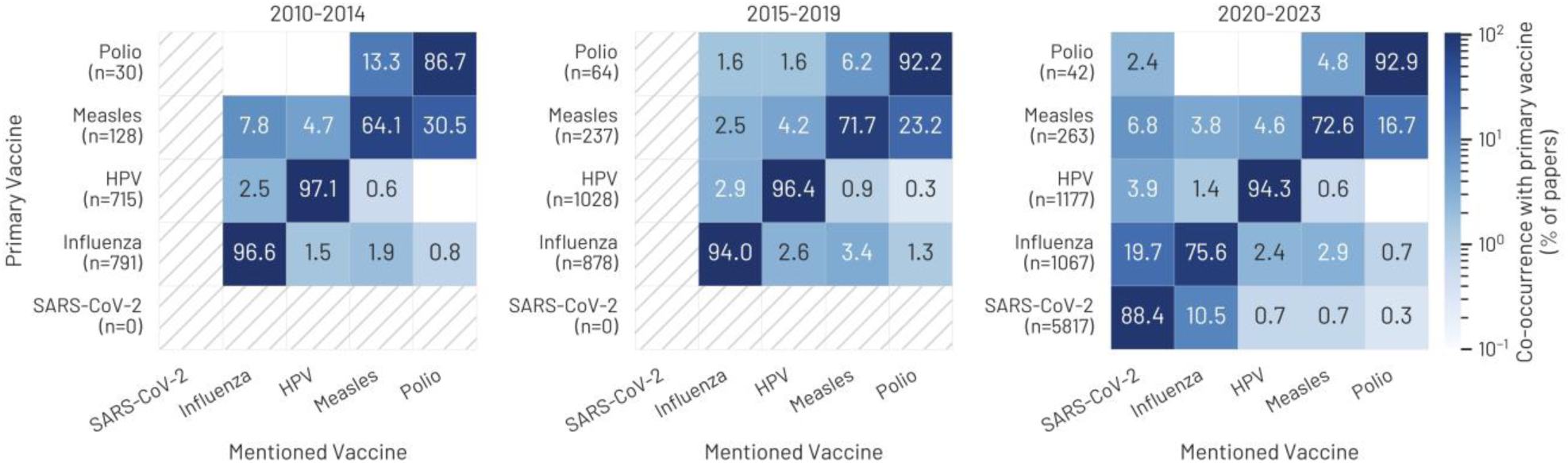
Co-occurrence of *primary vaccines* and *mentioned vaccines* from 2010 onwards. For each 5-year interval, the color-coded heatmap shows the percentage of articles focusing on a primary vaccine (y-axis) and mentioning another vaccine (x-axis). Percentages on the diagonal represent the proportion of papers that focus exclusively on a *primary vaccine*. Articles mentioning more than one additional vaccine are included more than once in the matrices. The total number of articles focusing on each *primary vaccine* for a given period is reported under the y-axis labels.

### Geographic patterns in global health research

We mapped the geographic distribution of global PSV research from 2010 onwards (Figure 3). To reconstruct the direction of research we evaluated country-specific proportions of publications authored by local institutions alone (*Domestic),* authored by or with institutions from a foreign country (*Inbound*), or focused exclusively on foreign countries (*Outbound)*.

**Figure 3.**
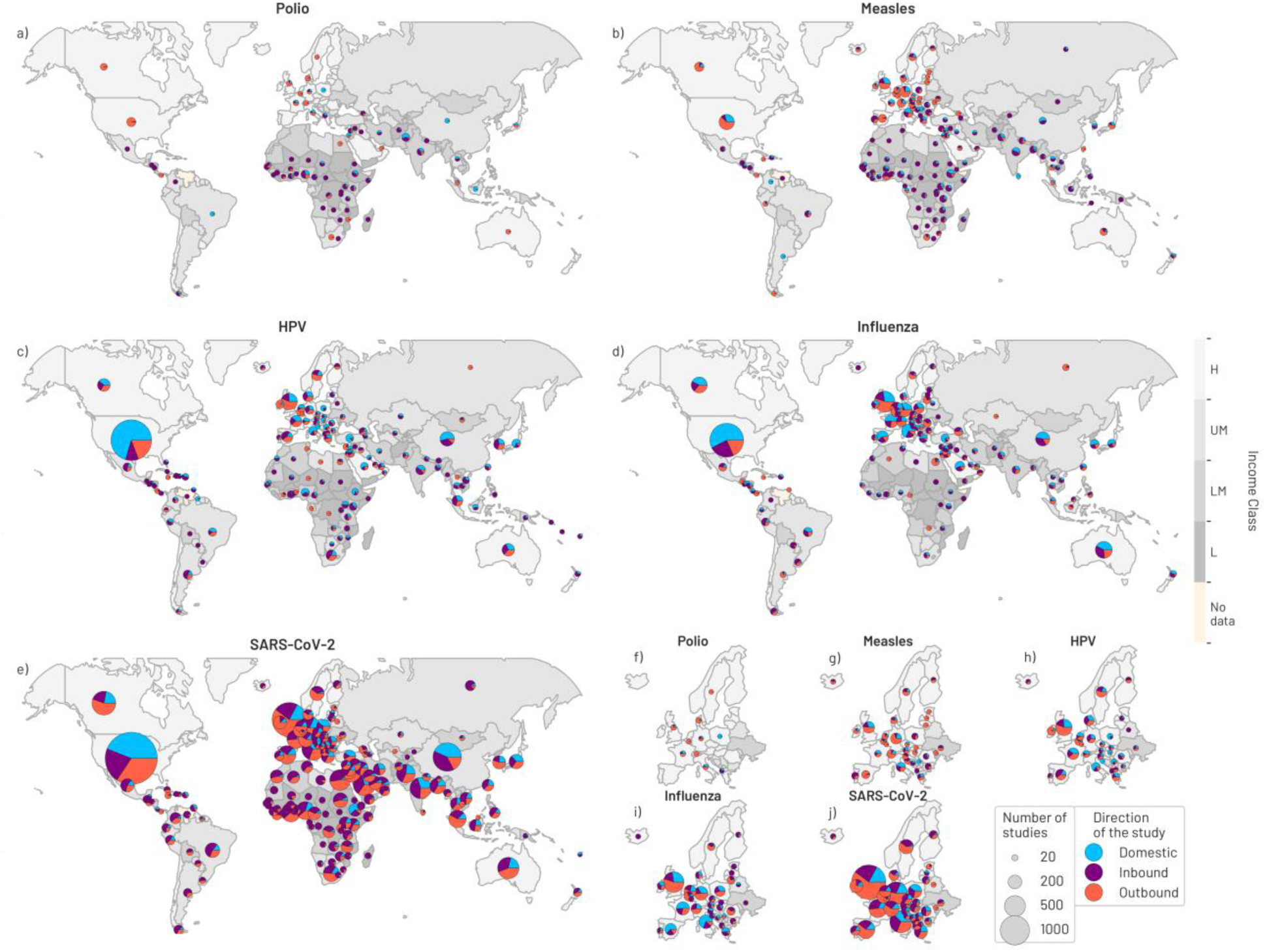
Geographic distribution of *author countries* and *studied countries* by vaccine from 2010 onwards. Pie slices represent country-specific shares of *Domestic*, *Inbound,* or *Outbound* studies. *Domestic*: studies on country *i* authored by local researchers alone. *Inbound*: studies on country *i* authored or co-authored by foreign institutions. *Outbound:* studies on any foreign country authored or co-authored by institutions from country *i*. The size of each pie chart is proportional to the total number of studies involving a country. Background colour corresponds to countries’ income levels. Panels *(f – j)* focus on European countries.

Overall, published PSV literature covered most countries worldwide, with HIC in North America and Europe authoring the majority of studies (72·2 %). However, notable differences emerged in the spatial distribution of published articles across the examined vaccines. PSV research on the polio vaccine was more geographically clustered compared to other vaccines, with hotspots of research in Sub-Saharan Africa, the Middle East, and South Asia. Conversely, research efforts on the influenza vaccine were relatively frequent in Europe and North America but scarce on the African continent, with over 70%, over 40%, and less than 30% of countries in the respective continents involved, *i.e.* authoring or being studied in at least one paper (<u>Figure S3, top row</u>).

We also observed income-related global imbalances in terms of research direction (Figure S3, bottom row). Despite the high number of studies on polio and measles vaccines authored by institutions in North America, Europe, and Australia, the direction of these studies was predominantly *Outbound* for HIC *(Median proportion Outbound studies, High-income; Polio: 82%; Measles: 61%)*, and mostly *Inbound* for low-income countries *(Median proportion Inbound studies, Low-income: Polio: 100%; Measles: 78%)*. In the same time frame, little *Domestic* and even less *Outbound* research emerged from low– and lower-middle-income countries. Similar but reversed income-dependent trends were observed in the distribution of PSV research on the HPV and influenza vaccines, although in HIC the shares of *Domestic*, *Inbound*, and *Outbound* research efforts were more equally distributed for these two vaccines *(Median proportion Domestic, Inbound, Outbound studies, High-income; Influenza: 24%, 37%, 26%; HPV: 33%, 32%, 33%)*. Lastly, we measured comparable shares of *Outbound* research on SARS-CoV-2 across income levels *(Median proportion Outbound studies SARS-CoV-2: Low-income: 21%; Low-middle-income: 31%; Upper-middle-income: 33%; High-income: 40%)*, with the amount of *Inbound* research decreasing with increasing country income, mirroring patterns observed for other vaccines.

### Drivers of global research flow

Through vaccine-specific logistic regression models, we assessed the potential factors influencing the probability of countries being the focus of global PSV research. The reconstructed marginal probabilities are shown in Figure 4 and Figure S4, upper panels, coefficients are reported in <u>Table S2 and Table S3</u>. We found that a significantly lower proportion of countries has been studied in polio research compared to all the other vaccines. For all vaccines, the probability of being studied was significantly associated with the country’s income level, although this dependence followed opposite directions for different vaccines. In the reference period (2010-2014), lower-income countries had a significantly higher probability of being the object of polio or measles PSV research compared to HIC. The disparity was greatest for the measles vaccine in the reference period, with HIC having 98% lower odds of being studied compared to the low-income group *(H vs L: OR=0·02; 95%CI = 0·00, 0·11)* (<u>Table S2)</u>. This income disparity remained constant over time for polio and appeared to level up for measles, with increased interest in higher-income settings but a decrease in lower-income ones. In contrast, HIC had a significantly higher probability of serving as study locations for the HPV and influenza vaccines, perpetuating existing income disparities without significant time-and-income variation. Additionally, the probability of being studied for the influenza vaccine appeared to be significantly higher in the years following the COVID-19 pandemic *(2020-23 vs 2010-14: OR=7·57; 95%CI = 1·16, 149·71)* (Table S3).

**Figure 4.**
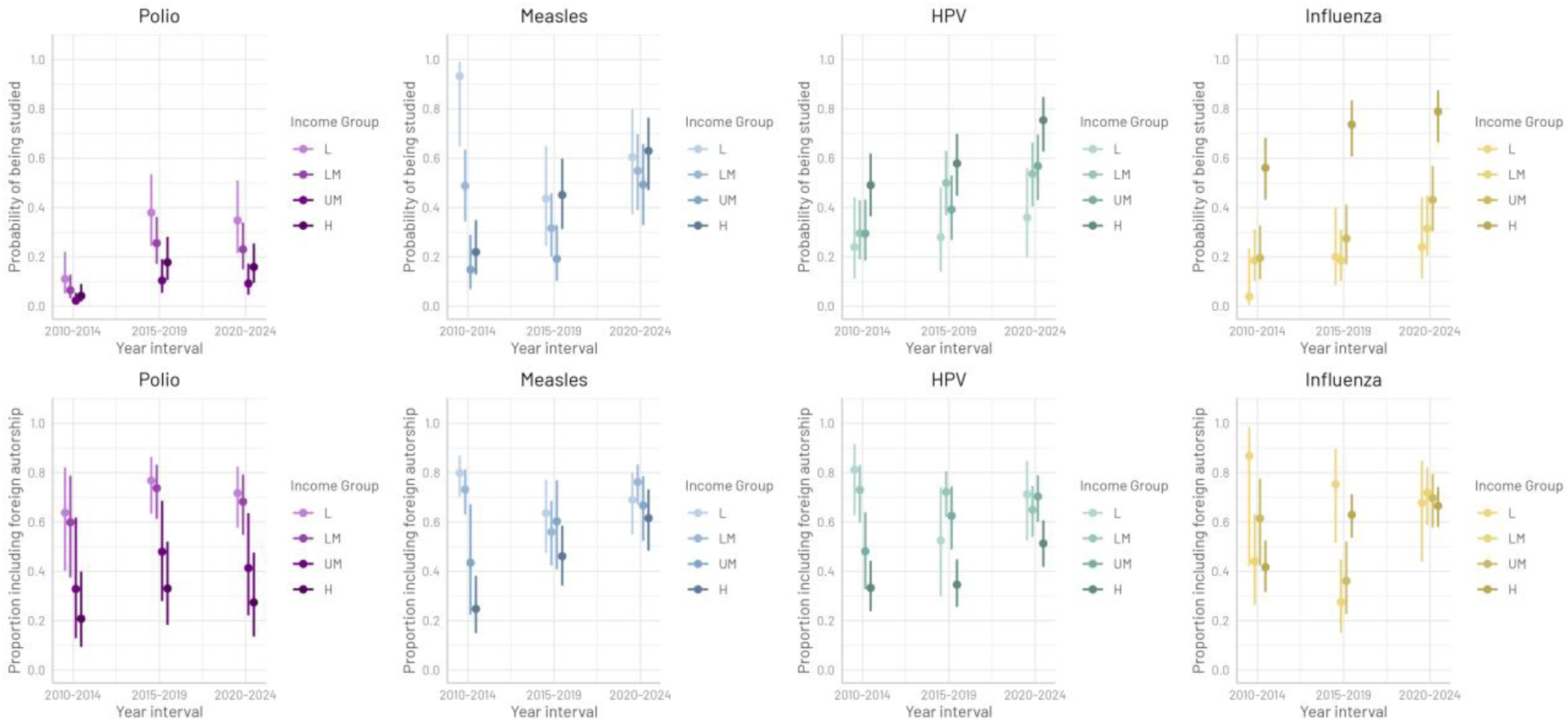
(Top row) Estimated probabilities, with 95% confidence intervals, of being the subject of PSV research on a given vaccine across time points and income groups. (Bottom row) Estimated proportion, with 95% confidence intervals, of PSV research articles on a given vaccine involving foreign researchers (*Inbound/(Inbound+Domestic)),* by income level of studied countries. L=low income; LM=lower-middle income; UM= upper-middle income; H= high income.

For both measles and polio, local disease incidence affected the probability of countries to be studied (Figure S4, <u>Table S2</u>). Countries where clinical polio cases were reported had 2·7 times higher odds of being studied compared to countries where the disease was eliminated *(Reported cases vs No cases: OR=2·67; 95%CI= 0·95, 7·37)*. Analogously, the probability of being studied was significantly higher among countries experiencing comparatively high measles incidence *(H vs L: OR=10·11; 95% CI= 4·45, 24·76)*. Conversely, we did not detect a consistent effect of national vaccination coverage on the probability of being studied for either vaccine.

### Determinants of foreign authorship

For countries that have been the object of PSV research, we modelled the proportion of articles (co-)authored by foreign countries using Beta regression models. The fitted values are shown in Figure 4 and Figure S4, bottom panels, the coefficients are reported in <u>Table S2 and Table S3</u>. Across all vaccines, during the reference period (2010-2014), the proportion of papers involving foreign research efforts increased as the income level of the *studied* country decreased. This income-related disparity appeared to diminish over time for the measles, HPV, and influenza vaccines. Conversely, we observed no significant time-dependent change for polio research.

For both polio and measles, countries reporting clinical disease cases had a smaller proportion of articles (co-)authored by foreign researchers. We found no consistent effect of national vaccination coverage on the proportion of foreign authorship.

## Discussion

This work examines the drivers of global health research using innovative methodologies, offering novel insights and analysis that, to our knowledge, are unexplored in the existing literature. We integrated an extensive literature search, machine learning-assisted article screening and data enrichment, network analysis, and statistical methods to develop a novel database of research papers on PSV, which offers a valuable resource for addressing broad global health research questions. Going beyond traditional bibliometric methods,^8,10–12^ and merging information on country-specific epidemiological and economic factors as well as direction of research, we investigated patterns and drivers shaping the global research efforts on PSV.

Our findings portrayed PSV research as a rather recent research field, partially shaped by vaccine-specific epidemiological conditions and unforeseen global health challenges, such as pandemic threats, but strongly influenced by country-specific economic determinants that steer academic focus and the opportunity for scientific collaborations.

By modelling the volumes of historical research data, we revealed that overall PSV publications have grown exponentially until 2020. Despite the eradication of polio and measled being within reach^25^, the research output on these vaccines remained relatively modest throughout the study period. Moreover, notwithstanding the rise in public vaccine hesitancy in recent years,^26,27^ it did not intensify as much as expected after COVID-19 pandemic, proving vulnerable to sudden disruptions. In contrast, we observed higher-than-expected post-pandemic publication volumes regarding the influenza vaccine, likely due to its analogies with the SARS-CoV-2 vaccines. These findings align with abundant evidence of the negative impact of the SARS-CoV-2 pandemic on routine childhood vaccinations, both in terms of vaccine uptake^28,29^ and public trust,^30^ and may be attributed to the temporary redirection of academic interest and research funding towards COVID-19-related research. The influence of SARS-CoV-2 on PSV research was further highlighted by a post-pandemic increase in articles on other *primary vaccines* mentioning the SARS-CoV-2 vaccine. This pattern, especially striking for vaccines that share few or no similarities with SARS-CoV-2 vaccine, likely reflects a momentary shift of the global health research agenda due to the very peculiar and worrisome situation of the pandemic threat. However, it also suggests that PSV research is highly sensitive to time– and context-dependent funding and publication opportunities, which in turn influence researchers’ actions.^16^ This result underscores the importance to maintain sustained academic attention on long-standing research objectives, such as vaccine-preventable diseases elimination and eradication efforts.

Our statistical analyses of geographical, economic, and epidemiological factors consistently showed that country income levels critically shape the landscape of PSV research, inducing an economically driven imbalance in both the global distribution and direction of research efforts.

In agreement with previous literature,^8,10–12^ we found that HIC produced the majority of global PSV research, typically redirecting their research on polio and measles towards LMIC and, on the other hand, attracting research efforts on HPV and influenza vaccines.

Our regression analysis also suggested a gradual shift in academic attention from LMIC to HIC, particularly for measles, but also for HPV and influenza. We showed that such a trend may be partially explained by changes in the epidemiology of the related diseases, although other factors likely played a significant role as well. These may include a country’s historical experience with the disease, the presence of local outbreaks, local vaccination recommendations and policies, political context, the unequal distribution of research funding, global publication bias, and countries’ involvement in international public health endeavours, such as disease elimination initiatives. These results also suggest that the gap left by shifting attention to HIC is not currently filled with *Domestic* research in LMIC.

Income-dependent disparities were also evident through the disproportionate amount of foreign authorship in studies concerning LMIC, especially in PSV research on polio and measles. This suggests a structural involvement of external researchers in LMIC, even in cases concerning very local health challenges. Except for polio research, where the historical imbalances persist, foreign research contributions became more evenly distributed across income groups for all vaccines, indicating an encouraging increase in academic cross-income-group collaborations. Combining this result with an overall increased attention to higher-income countries, however, our analyses also suggest that the levelling of foreign efforts might be driven by increased collaborations among HIC, likely at the expense of lower-income ones, where such research is often most needed.

Overall, our regression analyses provided strong evidence that a country’s lower income significantly limits its capacity to perform *Domestic* research, even in areas with high disease incidence, resulting in an inequitable and likely inefficient reliance on external research activities to address critical local health challenges.

We acknowledge some limitations. First, our literature search strategy primarily employed English-language keywords and a substantial portion of non-English scientific literature may not be indexed in the two databases used. As a result, we may have missed relevant studies published in other languages lacking an English abstract. Because English-language publications are not fully representative of the global scientific landscape, this may have introduced a language bias into our literature corpus. However, the vast majority of global scientific output is currently published in English.^31^ Hence, any language bias is likely to be limited in scope and not expected to substantially affect the main patterns identified in the analysis.

Second, although our ML-assisted article selection achieved strong performance metrics, some misclassification may have occurred, potentially leading to the inclusion of a few irrelevant articles or the exclusion of some pertinent ones. Yet, given the scale of the dataset and the inherent subjectivity in relevance judgements, even a fully manual screening process would likely have introduced comparable inconsistencies. Importantly, the volume and the consistency of the retained articles still provide a reliable basis for mapping broader scientific trends.

Lastly, inclusion of additional contextual factors that may shape global attention on PSV research could offer deeper explanatory power. However, in order to ensure complete, consistent and reliable cross-country comparison on a global scale, we chose to leverage only indicators provided by internationally recognised organizations. Collating national datasets may have introduced uncertainty and unknown sources of potential bias, ultimately compromising the robustness and comparability of our analysis.

## Conclusion

The persistent misalignment between countries with the financial capacity to conduct PSV research and those most in need of its outcomes highlights entrenched inequities within this field. These disparities are not unique to PSV research but are rather a symptomatic of broader structural imbalances that pervade global health research. Research priorities –particularly in HIC-are often shaped by internal agendas and funding opportunities rather than global need. This hampers the potential impact of research efforts in achieving broader health outcomes, such as increased vaccine uptake in underserved settings.^32^

Our findings suggest that such imbalances are likely to be exacerbated during periods of global disruptions, as demonstrated by the COVID-19 pandemic, with potentially detrimental consequences for both ongoing public health interventions and future research directions. By providing a clear view of the main drivers of PSV research and revealing the risks of shifting academic focus, this study highlights the need to decolonise PSV research and, broadly, global health research. Addressing historical imbalances requires promoting equitable, inclusive research partnerships, and redistributing leadership, ownership, and priority-setting power across the global research community. Key principles for establishing equitable research dynamics include sustainability, policy-oriented objectives, transparency, equitable data access, and meaningful community engagement.^33–37^ Existing ethical guidance can help researchers design projects in alignment with these values,^38^ and funding mechanisms should embed strong incentives to promote such practices.^39^ Future funding allocation criteria should prioritise research with the highest potential health impact for vulnerable populations – such as improving vaccine coverage among under-immunised children – over purely academic or innovation-driven considerations.^40^ Grants must be accessible to institutions in LMIC directly, fostering application diversity and enhancing local research capacity.^3,41^ Achieving key global health targets, such as the eradication of polio and measles, requires sustainable investments guided by clear long-term strategies and implemented within well-defined timeframes, particularly in low-resource settings.^42^ Currently, decision-making power remains highly concentrated in a small group of actors, mostly from HIC, yet continued support may be subject to political or economic fluctuations. This can lead to funding disruptions with potentially serious implications for the populations affected. A sustainable transition toward increased domestic investment in health research is therefore critical and must include robust exit strategies, early planning, stakeholder engagement, and dedicated pre-transition resources. ^3,43,44^ Scientific journals should also play a crucial role in reshaping inequitable research practices by establishing systems to identify and discourage submissions that violate equity principles related to content and authorship.^45,46^ Our work highlights the need for a systemic change across funding structures, authorship norms, and publication practices in order to redress long-standing inequities in global health research, and to ensure that research efforts ultimately benefit the most affected.^47^

## Data availability statement

The database containing all scientific publications included in this analysis will be made available on reasonable request for scientific research purposes. For further information or to request access, please contact the corresponding author.

## Patient and Public Involvement statement

It was not appropriate or possible to involve patients or the public in the design, or conduct, or reporting, or dissemination plans of our research

## Author contributions

DB, VO, ZA, CC, LPL, AT, SS, JAP, and AM contributed to the conceptualization by formulating the research idea, objectives, and research questions. VO, LPL, ED, and AM developed the search string, screening strategy, and annotated the dataset for relevance. DS and JAP collected and curated the dataset, while DB, VO, ZA, and CC enriched it with relevant attributes and performed data cleaning. DB, VO, ZA, CC, and AM designed the study methods and performed technical analyses, including machine-assisted literature searches, enrichment via natural language processing, network analysis, and other formal analyses. CC and ZA conducted the regression analysis. DB, VO, ZA, CC, and AM produced the results and drafted the initial manuscript. All authors interpreted the results, reviewed, and edited the drafts. DB and VO led and coordinated the project, AM supervised and secured funding.

