## Supplement for "Mapping Inequities in Global Vaccine Sentiment Research"

### Supplementary information for: Mapping Inequities in Global Vaccine Sentiment Research

#### *Methodological details*

##### Location extraction

First, using a pre-trained large language model, *GPT-3.5-turbo,* via the OpenAI APIs, we inferred the geographical entities corresponding to the countries where a study was performed. We provided *GPT-3.5-turbo* with a custom prompt, shown in the following, as well as the title and the abstracts of each research article, asking for the extraction of any geographical entity, returned as the standardized ISO 3166-1 alpha-3 codes of the corresponding countries:

*"The following text might contain a geographical location or multiple geographical locations: {text}. Do not output the country from mentions to languages. Output only the standard ISO English name of the countries and the ISO 3166-1 alpha-3 codes, structured as JSON like in the following: { "countries": [ { "name": "Hong Kong", "iso_code": "HKG" }, { "name": "Ghana", "iso_code": "GHA" } ] }. Output 'no_country' and 'NOCODE' if no country is matched. Result: "*

Second, screening the affiliation of the authors with a similar technique, we inferred the geographical entities corresponding to the countries performing a study. To ensure maximal precision, this task required a preprocessing step. We first separated each institution from the list of institutions participating in each study. Where possible, we directly extracted the country of affiliation using regular expression. We then retained the institutions which were not successfully mapped, and screened them using *GPT-3.5-turbo* with the following custom prompt:

*"The following text is the name of a research institution, which might contain a geographical location or multiple geographical locations: {text}. Your purpose is to extract the geographical locations at country level. Do not output the country from mentions to languages. Output only the*

*standardized name of the institution, removing departments or similar entities, the standard ISO English name of the countries and the ISO 3166-1 alpha-3 codes, structured as JSON like in the following: {{ "countries": [ {{ "inst": "Hong Kong University" "name": "Hong Kong", "iso_code": "HKG" }}, {{ "inst": "New York University (NYU)"" "name": "United States", "iso_code": "USA" }} ] }}. Output 'no_country' and 'NOCODE' if no country is matched. Result: ”*

Thanks to the contextual flexibility that Large Language Models can provide, this approach allowed us to easily map studies performed in specific regions to their respective countries and to infer the location of institutions.

##### Country income, vaccine coverage, and disease incidence

We assigned to each country a list of categorical attributes, *i.e.* income level,[3] disease incidence for each related disease,[4,5] and vaccine-specific coverage over time.[6,7] We categorized countries into four income level groups (*L=low; LM=lower-middle; UM= upper-middle; H= high*) according to the World Bank’s Gross National Income (GNI) per capita as of 2021.[3] The income level for each country was fixed throughout time intervals.

Attributes regarding incidence and coverage were defined for measles and polio, as information regarding HPV and Influenza was incomplete for a considerable number of countries. We calculated the mean vaccination coverage for each country within specified 5-year intervals between 2000 and 2023. Then, we classified each country as *L=low, LM=lower-middle, UM= upper-middle,* or *H= high* coverage, according to the quartiles of the global distribution of vaccination coverage in that 5-year interval. The same procedure was applied to incidence data computing the mean yearly incidence of disease (cases per 100,000 residents), in each 5-year interval for each country, and attributing incidence levels based on quartiles. For polio incidence, countries were classified as "0 cases" or “>0 cases”.

#### Trends analysis

We modelled the volume of published papers for each vaccine in recent years, assuming exponential growth. We chose to conduct this analysis from 2010 onwards in order to include the licensing of the HPV and 2009/H1N1 pandemic influenza vaccines. We fitted the yearly number of published papers *Y* from 2010 to 2019, the pre-COVID-19-pandemic years, with an exponential curve

$$Y = \alpha * e^{\beta * t}$$

where $\alpha$ and $\beta$ are coefficients to be estimated and *t* is a given year. All fits achieved an *Adjusted-* $r^2 > 0.6$. For each vaccine, we compared $y_t^{true}$ , the measured volume of publications for the years 2020–2023, with the expected number of publications predicted by the fitted models $y_t^{pred}$ in the same period, computing the *Percentage Difference* of the total volumes of publication

$$\text{Percentage Difference} = \left(\frac{\sum_t y_t^{true} - \sum_t y_t^{pred}}{\sum_t y_t^{pred}}\right) \times 100$$

The confidence intervals of the *Percentage Difference* were evaluated by comparing the measured volume of publications with the 95%CI of the prediction, i.e. by substituting $y_t^{pred}$ with the lower and upper bounds of the predictions.

#### Statistical Analysis

We modelled the probability of a country to be *studied*, with a logistic regression model for each vaccine. Let Y be the binary dependent variable indicating whether each country had been the subject of at least one research paper on a given vaccine *(1 if studied, 0 if not)* in each time period. For countries $i = 1, \ldots, n$ and covariates $X_1, \ldots, X_m$ we assume $Y_i | \; x_1, \ldots, x_m \sim Ber(p_i)$. The expected probability of each country being studied $E[Y_i | x_1, \ldots, x_m|] = p_i$ is then specified as follows

$$\log i\,t(p_i) \;=\; \beta_0 + \beta_1(TimeInterval_i) + \beta_2(IncomeLevel_i)$$
$$+\; \beta_3(IncomeLevel_i \times TimeInterval_i) + \beta_4\left(VaccinationCovergae_{i,t}\right)$$
$$+\, \beta_5\left(DeasesIncidence_{i,t}\right)$$

where $\beta_0$ is the intercept, $\beta_1$, $\beta_2$, $\beta_3$, $\beta_4$ and $\beta_5$ are the regression coefficients for the covariates. Each covariate's lowest level (2010-2014 for time, "Low" for income, vaccination coverage, and disease incidence) was taken as a reference level to estimate coefficients.

We further modelled the proportion of papers authored in collaboration with, or exclusively by, foreign researchers, evaluated as the number of *Inbound* studies conducted in a given country over the total number of studies conducted there (*Inbound* + *Domestic).* Let $Y$ be the random variable describing the proportion of papers authored with or by foreign authors in each country; we assume $Y_i \mid x_1, \; \ldots, \; x_m \sim Beta(\mu_i, \; \phi) \mid$ , where $E(Y_i \mid x_1, \; \ldots \;, \; x_m \mid) \;=\; \eta_i$ and $\mathrm{Var}(Y_i \mid x_1, \; \ldots \;, \; x_m) \;=\; \frac{\mu_i(1-\mu_i)}{(1+\phi)}$ . The parameter $\phi$ is known as the precision parameter. The Beta regression model is defined as

$$\log i\,t(\mu_i) \;=\; \gamma_0 + \gamma_1(TimeInterval_i) + \gamma_2(IncomeLevel_i)$$
$$+\; \gamma_3(IncomeLevel_i \times TimeInterval_i) + \gamma_4\left(VaccinationCovergae_{i,t}\right)$$
$$+\, \gamma_5\left(DeasesIncidence_{i,t}\right)$$

Both regression models were fitted using maximum likelihood estimation. The significance of the coefficients was tested using Wald tests, and statistical significance was set to $\alpha$ *=0.95*. All the statistical analyses were performed using *R* software.

As outputs of the regression models, we reported the complete tables of coefficients, and we plotted the estimated probabilities (Table S2, Table S3)

#### *Supplementary Tables*

**Table S1** Literature search string used to retrieve scientific articles from the MEDLINE and Web of Science Core Collection databases without geographic or time constraints. The column "Field" indicates the section of the article in which the search term had to be mentioned for the paper to be captured by the string.

| **String component** | **Search terms** | **Field** |
|---|---|---|
| 1 | immunis* OR immuniz* OR vaccin* OR vax OR jab OR jabs | Title |
| 2 | "mis-trust*" OR mistrust* OR "dis-trust*" OR distrust* OR "mis-conce*" OR misconce* OR "anti-vax*" OR antivax* OR "anti-vaccin*" OR antivaccin* OR "mis-belie*" OR misbelie* OR hesita* OR refus* OR resistan* OR reject* OR reluctan* OR vacillat* OR skeptic* OR sceptic* OR concern* OR trust* OR accept* OR agree* OR confiden* OR complian* OR adheren* OR aware* OR willing* OR barrier* OR fear* OR anxiet* OR exemption* OR oppos* OR objection* OR object* OR delay* OR dropout* OR controvers* OR attitud* OR perspective* OR view* OR choice* OR belie* OR doubt* OR opinion* OR knowledge OR perceiv* OR percept* OR motivat* OR sentiment* OR emotion* OR rumor* OR rumour* OR uptake OR intent* OR decis* OR indecis* OR uncertain* OR behave* OR dilemma* | Title or abstract |
| 3 | vaccinia | Title or abstract |
| **Full string** | **1 AND 2 NOT 3** | |

**Table S2** Coefficients of the logistic regression models (#), shown as Odds Ratios, assessing factors influencing the likelihood of a country to be the object of studies on polio or measles vaccines, and of the Beta regression models (§) assessing which country attributes influenced the proportion of PSV research papers on a given vaccine (polio or measles) (co-)authored by foreign countries. In (#) the dependent outcome was a binary variable indicating whether a country had been the subject of at least one PSV research paper on a given vaccine (polio or measles). In (§) the outcome variable was evaluated as the number of *Inbound* studies conducted in a given country over the total number of studies conducted there (*Inbound* + *Domestic*). For both models, independent variables included time intervals, country income level, vaccination coverage, and disease incidence. L=low; LM=lower-middle; UM= upper-middle; H= high. The table reports the Odds Ratios and the 95% confidence intervals.

| | **Polio** | | **Measles** | |
|---|---|---|---|---|
| | **Likelihood to be studied[#]** | **Proportion foreign authorship[§]** | **Likelihood to be studied[#]** | **Proportion foreign authorship[§]** |
| ***Time interval (ref: 2010-2014)*** | | | | |
| **2015-2019** | 4.91<br>(2.37, 11.09) | 1.88<br>(0.76, 4.61) | 0.05<br>(0.00, 0.33) | 0.44<br>(0.19, 0.97) |
| **2020-2024** | 4.29<br>(2.02, 9.86) | 1.43<br>(0.54, 3.77) | 0.10<br>(0.00, 0.70) | 0.55<br>(0.26, 1.18) |
| ***Income level (ref: L)*** | | | | |
| **LM** | 0.56<br>(0.29, 1.09) | 0.84<br>(0.43, 1.65) | 0.06<br>(0.00, 0.37) | 0.68<br>(0.35, 1.32) |
| **UM** | 0.19<br>(0.07, 0.44) | 0.27<br>(0.09, 0.77) | 0.01<br>(0.00, 0.07) | 0.19<br>(0.06, 0.58) |
| **H** | 0.35<br>(0.15, 0.81) | 0.14<br>(0.05, 0.42) | 0.02<br>(0.00, 0.11) | 0.08<br>(0.03, 0.19) |
| ***Coverage level (ref: L)*** | | | | |
| **LM** | 0.94<br>(0.49, 1.81) | 1.38<br>(0.71, 2.67) | 2.12<br>(1.83, 3.87) | 0.97<br>(0.64, 1.49) |
| **UM** | 0.90<br>(0.40, 1.97) | 0.76<br>(0.29, 2.00) | 1.07<br>(0.56, 2.05) | 0.93<br>(0.58, 2.17) |
| **H** | 0.62 | 0.61 | 0.56 | 1.12 |

| | | | | |
|---|---|---|---|---|
| | (0.23, 1.54) | (0.21, 1.79) | (0.26, 1.17) | (0.58, 2.17) |
| ***Incidence level (ref polio: No cases; ref measles: L)*** | | | | |
| **Polio: Reported cases/ measles: LM** | 2.67<br>(0.95, 7.37) | 0.41<br>(0.15, 1.14) | 9.18<br>(4.15, 21.75) | 0.41<br>(0.17, 0.96) |
| **Measles: UM** | na | | 12.80<br>(5.90, 30.16) | 0.65<br>(0.28, 1.49) |
| **Measles: H** | na | | 10.11<br>(4.45, 24.76) | 0.70<br>(0.29, 1.66) |
| **Measles: Missing** | na | | 1.32<br>(0.45, 3.72) | 0.37<br>(0.11, 1.24) |
| ***Time interval & income level (refs: 2010-2014; L)*** | | | | |
| **2015-2019 & LM** | na | | 8.73<br>(1.16, 184.43) | 1.05<br>(0.36, 3.05) |
| **2015-2019 & UM** | | | 24.63<br>(2.88, 555.96) | 4.48<br>(1.02, 19.64) |
| **2015-2019 & H** | | | 52.67<br>(7.05, 1114.71) | 5.90<br>(1.94, 17.96) |
| **2020-2024 & LM** | | | 11.65<br>(1.47, 251.24) | 2.10<br>(0.79, 5.52) |
| **2020-2024 & UM** | | | 50.94<br>(5.77, 1165.23) | 4.65<br>(1.18, 18.29) |
| **2020-2024 & H** | | | 55.19<br>(6.79, 1206.75) | 8.69<br>(2.82, 26.72) |
| **Constant** | 2.008 | 0.658 | 0.684 | 1.888 |
| **Observations** | 543 | 84 | 561 | 252 |
| **R2** | | 0.414 | | 0.259 |
| **Log Likelihood** | -208.112 | 154.232 | -294.939 | 308.408 |

**Table S3** Coefficients of the logistic regression models (#), shown as Odds Ratios, assessing factors influencing the likelihood of a country to be the object of studies on the HPV or influenza vaccines, and of the Beta regression models (§) assessing which country attributes influenced the proportion of PSV research papers on a given vaccine (HPV or influenza) (co-)authored by foreign countries. In (#) the dependent outcome was a binary variable indicating whether a country had been the subject of at least one PSV research paper on a given vaccine (polio or measles). In (§) the outcome variable was evaluated as the number of *Inbound* studies conducted in a given country over the total number of studies conducted there (*Inbound* + *Domestic*). For both models, independent variables included time intervals and country income level. L=low; LM=lower-middle; UM= upper-middle; H= high; HPV= human papilloma virus.

| | **HPV** | | **Influenza** | |
|---|---|---|---|---|
| | **Likelihood to be studied#** | **Proportion foreign authorship§** | **Likelihood to be studied#** | **Proportion foreign authorship§** |
| ***Time interval (ref: 2010-2014)*** | | | | |
| **2015-2019** | 1.23 (0.34, 4.50) | 0.25 (0.06, 0.97) | 5.99 (0.87, 120.03) | 0.46 (0.04, 5.29) |
| **2020-2024** | 1.78 (0.52, 6.35) | 0.57 (0.16, 1.96) | 7.57 (1.15, 149.71) | 0.32 (0.02, 3.55) |
| ***Income level (ref: L)*** | | | | |
| **LM** | 1.33 (0.46, 4.21) | 0.62 (0.20, 1.90) | 5.45 (0.95, 103.13) | 0.12 (0.01, 1.23) |
| **UM** | 1.31 (0.45, 4.20) | 0.21 (0.06, 0.67) | 5.85 (1.02, 110.76) | 0.24 (0.02, 2.49) |
| **H** | 3.05 (1.11, 9.42) | 0.11 (0.04, 0.33) | 30.71 (5.85, 567.91) | 0.10 (0.01, 1.02) |
| ***Time interval & income level (refs: 2010-2014; L)*** | | | | |
| **2015-2019 & LM** | 1.92 (0.42, 8.64) | 3.73 (0.80, 17.26) | 0.16 (0.00, 1.46) | 1.03 (0.07, 14.85) |
| **2015-2019 & UM** | 1.25 (0.27, 5.72) | 6.98 (1.42, 34.16) | 0.25 (0.01, 2.23) | 0.76 (0.05, 10.58) |
| **2015-2019 & H** | 1.15 (0.26, 5.03) | 4.13 (1.02, 19.18) | 0.36 (0.01, 2.96) | 5.09 (0.41, 62.12) |
| **2020-2024 & LM** | 1.54 (0.34, 6.63) | 1.188 (0.28, 5.01) | 0.26 (0.01, 2.18) | 10.12 (0.75, 136.14) |

| | | | | |
|---|---|---|---|---|
| **2020-2024 & UM** | 1.77<br>(0.39, 7.73) | 4.44<br>(1.02, 19.18) | 0.41<br>(0.01, 3.35) | 4.48<br>(0.34, 59.08) |
| **2020-2024 & H** | 1.78<br>(0.40, 7.68) | 3.66<br>(0.93, 14.44) | 0.38<br>(0.01, 3.06) | 8.67<br>(0.73, 103.11) |
| **Constant** | 1.15 | 1.46 | 3.18 | 1.88 |
| **Observations** | 561 | 262 | 561 | 214 |
| **R2** | | 0.197 | | 0.164 |
| **Log Likelihood** | -362.45 | 192.6 | -307.21 | 129.3 |

#### *Supplementary Figures*

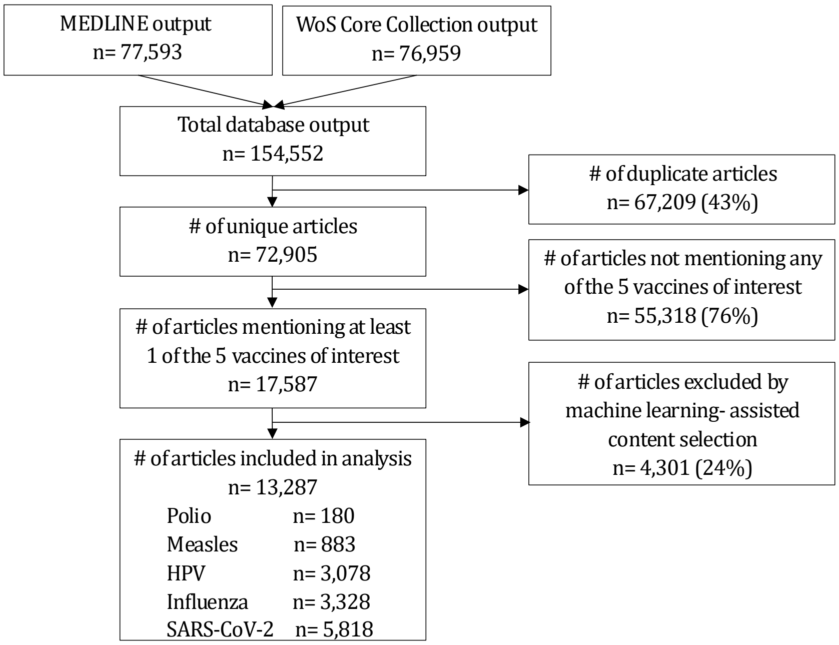


**Figure S1** Overview of the literature search and screening process. SARS-CoV-2= Severe Acute Respiratory Syndrome CoronaVirus 2; HPV= Human papillomavirus

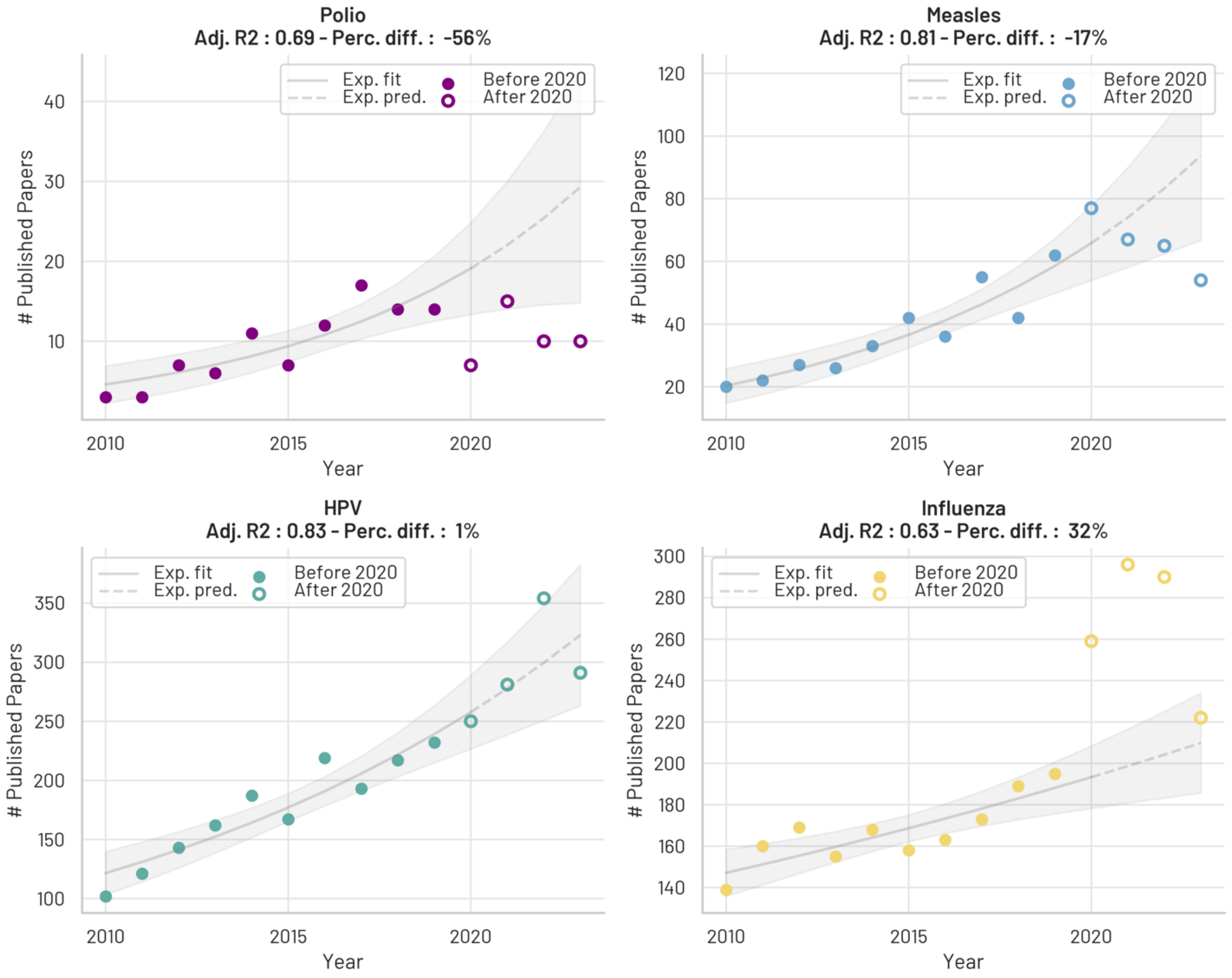


**Figure S2** Temporal trend of publication volumes for PVS literature on each *primary vaccine* (excluding SARS-CoV-2), modelled via exponential curves. The dots show the observed number of published articles by year for a given *primary vaccine*, before (filled) and after (hollow) the emergence of the SARS-CoV-2 pandemic (2020). In each subplot, the solid line shows the exponential curve fitted to the observed data before 2020, and the dashed line reports the predicted number of publications in pandemic years. The shaded areas report the 95% confidence interval of the fitted and predicted curves. The Adjusted-R2 and the Percentage difference (the excess proportion in the observed annual number of articles compared to the predictions) are reported in the titles.

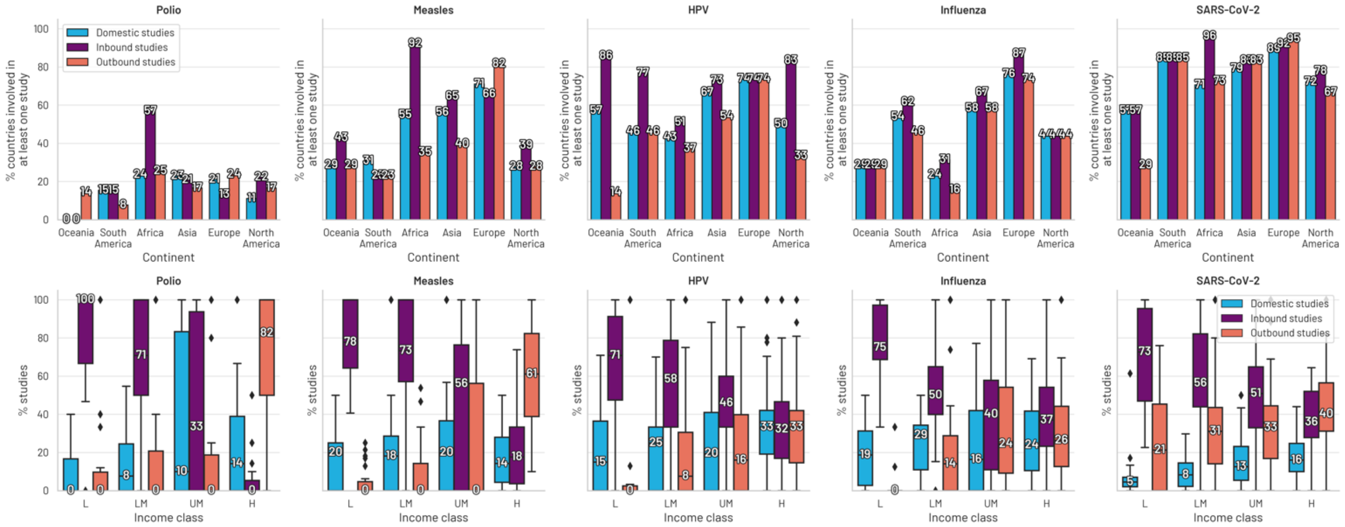


**Figure S3** Top row: Proportion of countries per continent involved in at least one Domestic (light blue), Outbound (orange), or Inbound (purple) PSV study for each vaccine. Bottom row: Boxplots showing the distribution of the share of Domestic, Inbound, or Outbound studies across countries grouped by income class (L = Low, LM = Lower-middle, UM = Upper-middle, H = High). The boxplot central lines and values represent the median proportion of studies for each income group.

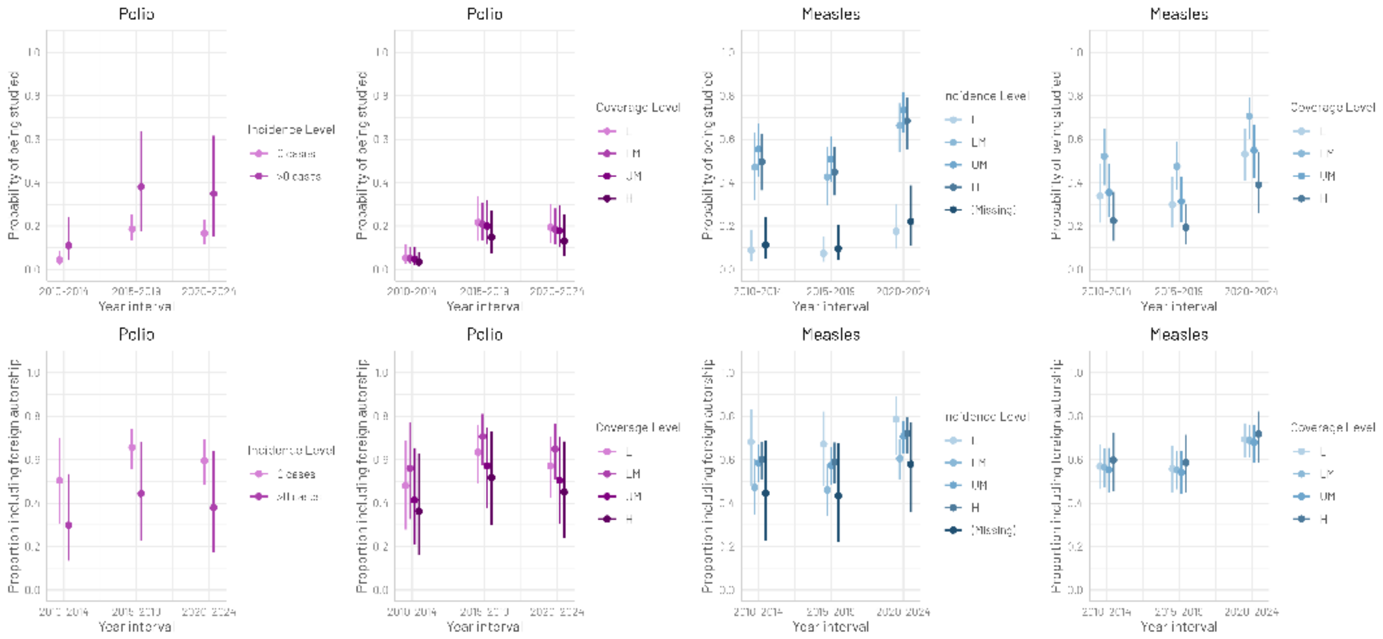


**Figure S4 Upper row:** Estimated probabilities, with 95% confidence intervals, of being the object of PVS research on a given vaccine across time points by levels of disease incidence and vaccination coverage. **Bottom row:** Estimated proportion, with 95% confidence intervals, of PSV research articles on a given vaccine involving foreign researchers over time and by levels of disease incidence and vaccination coverage.

3 World Bank Group. World Bank Country and Lending Groups. 2024.

4 World Health Organization. Poliomyelitis - number of reported cases. 2024.https://www.who.int/data/gho/data/indicators/indicator-details/GHO/poliomyelitis---number-of-reported-cases (accessed 15 Jul2024).

5 WOrld Health Organization. Measles - number of reported cases. 2024.https://www.who.int/data/gho/data/indicators/indicator-details/GHO/measles---number-of-reported-cases (accessed 15 Jul2024).

6 World Health Organization. Polio (Pol3) Immunization coverage estimates by country. 2024.https://apps.who.int/gho/data/node.main.A831 (accessed 15 Jul2024).

7 World Health Organization. Measles-containing-vaccine second-dose (MCV2) immunization coverage by the locally recommended age (%). 2024.https://www.who.int/data/gho/data/indicators/indicator-details/GHO/measles-containing-vaccine-second-dose-(mcv2)-immunization-coverage-by-the-nationally-recommended-age-(-) (accessed 15 Jul2024).
